# Genomic Risk Scores and Oral Contraceptive-Associated Ischemic Stroke Risk

**DOI:** 10.1101/2022.09.29.22280517

**Authors:** Forrest Lin, Liisa Tomppo, Brady Gaynor, Kathleen Ryan, John W. Cole, Braxton D. Mitchell, Jukka Putaala, Steven J. Kittner

**Affiliations:** Department of Neurology, Baltimore Veterans Affairs Medical Center and University of Maryland, School of Medicine, Baltimore, MD, USA; Department of Neurology, Helsinki University Hospital and University of Helsinki, Helsinki, Finland; Department of Medicine, University of Maryland, School of Medicine, Baltimore, MD, USA; Geriatrics Research and Education Clinical Center, Baltimore Veterans Administration Medical Center, Baltimore, MD

## Abstract

**Background:** Oral contraceptives (OCs) are generally safe but vascular risk factors increase OC-associated ischemic stroke risk. We performed a case-control study to evaluate whether a genomic risk score for ischemic stroke modifies OC-associated ischemic stroke risk.

**Methods:** The Genetics of Early-Onset Stroke (GEOS) study includes 340 premenopausal women (143 ischemic stroke cases and 197 controls) with data on OC use within 30 days before the index event (for cases) or interview (for controls). Using a previously validated genetic risk score (metaGRS) for ischemic stroke based on 19 polygenic risk scores of vascular events and risk factors, we stratified our sample into tertiles of genomic risk. We evaluated the association between OC use and ischemic stroke within each tertile. We tested if the association between OC use and ischemic stroke depended on the genomic risk of stroke using logistic regression with an OC use × metaGRS interaction term.

**Results:** Among all women, OC use was significantly associated with ischemic stroke (odds ratio = 2.4, p = 0.002). The odds ratio for ischemic stroke associated with OC use increased from 1.5 in the tertile with the lowest genomic risk to 3.9 in the tertile with the highest genomic risk of ischemic stroke. The formal test of interaction was consistent with our hypothesis (p = 0.07) that the genomic risk score modifies the association of OC use with ischemic stroke.

**Conclusions:** Our results suggest that genomic profile modifies the OC-associated ischemic stroke risk. Larger studies are warranted to determine whether a genomic risk score could be clinically useful in reducing OC-associated ischemic stroke.

## Introduction

Combined oral contraceptives (OCs) are associated with an increased risk of ischemic stroke.^1–3^ Although ischemic strokes are rare in reproductive-aged women, the consequences are often severe. Despite several factors that increase OC-associated ischemic stroke have been identified, including age, smoking, and hypertension,^4^ the exact mechanisms remain insufficiently understood. Identifying those women at risk is important to reduce the OC-associated risk of ischemic stroke. The aim of our study was to evaluate whether genetic predisposition for ischemic stroke, measured by a validated genomic risk score, modifies OC-associated ischemic stroke risk.

## Methods

### Study cohort

The Genetics of Early Onset Stroke (GEOS) Study is a population-based case-control study designed to identify genetic determinants of early-onset ischemic stroke and to characterize interactions with environmental risk factors. Stroke cases aged 15-49 years old at the time of ischemic stroke were recruited from the greater Baltimore-Washington area over 4 time periods between 1992–2008, along with age and sex matched controls.^5^ Study participants provided information from a standardized interview on age, history of hypertension, diabetes, and coronary artery disease. Current smoking status and OC use were defined use within 1 month before the event for cases and at a comparable reference time for controls.^5^ The study was approved by the University of Maryland at Baltimore Institutional Review Board, and all participants gave written informed consent.

The present study was limited to 340 premenopausal women of European ancestry with available data on OC use. Only participants of European ancestry were included because the genomic risk score was derived and validated in this population.

### Genomic risk score and analysis

Study subjects were genotyped with the Illumina 1M array and imputed using the TOPMed reference panel on the University of Michigan Imputation Server, as previously described.^6^

We used a previously validated genomic risk score (metaGRS) for ischemic stroke that is based on 19 polygenic risk scores for vascular events and cardiovascular risk factors and validated in nearly 400,000 subjects from the UK BioBank.^7^ We first stratified our sample of GEOS women into tertiles of genomic risk based on their metaGRS scores and evaluated the association between OC use and ischemic stroke within each tertile. We then tested if the association between OC use and ischemic stroke depended on genomic risk of stroke using logistic regression with an OC use × metaGRS interaction term.

### Data sharing

The aggregated data that support the findings are available by corresponding author upon reasonable request.

## Results

Characteristics of the 143 cases and 197 controls are shown in Table 1. Cases were more likely to report OC use than controls (25% vs 12%; OR = 2.4 (95% C.I: 1.4-4.3); p = 0.002). Cases were also more likely to smoke (45.5% vs 21.8%, p = < 0.001) and to report a history of diabetes (15.0% vs 7.8%; p = 0.06), although there was little difference between the groups in other comorbidities.

**Table 1.**
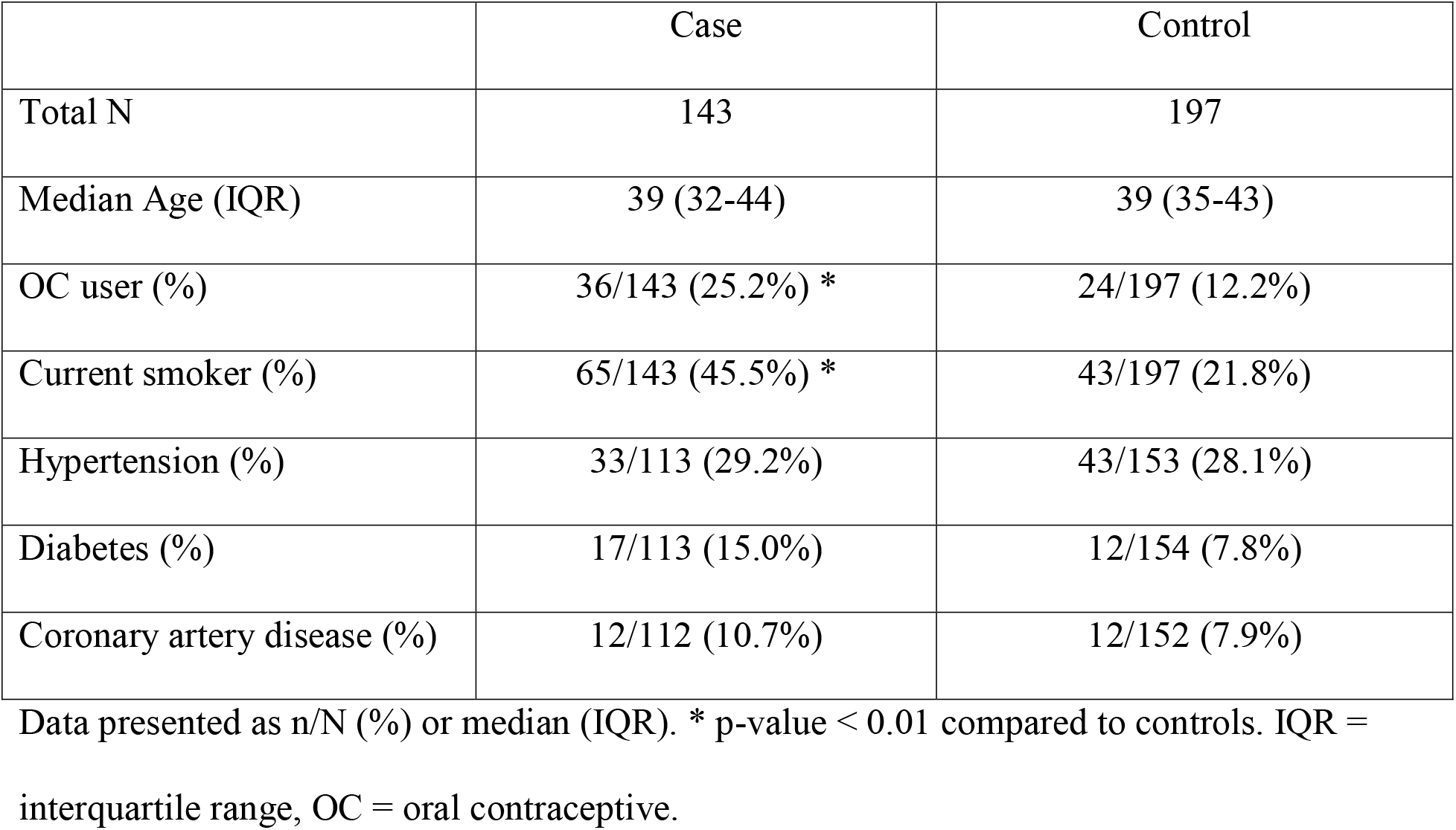
Basic characteristics according to case control status

|  | Case | Control |
| --- | --- | --- |
| Total N | 143 | 197 |
| Median Age (IQR) | 39 (32-44) | 39 (35-43) |
| OC user (%) | 36/143 (25.2%) * | 24/197 (12.2%) |
| Current smoker (%) | 65/143 (45.5%) * | 43/197 (21.8%) |
| Hypertension (%) | 33/113 (29.2%) | 43/153 (28.1%) |
| Diabetes (%) | 17/113 (15.0%) | 12/154 (7.8%) |
| Coronary artery disease (%) | 12/112 (10.7%) | 12/152 (7.9%) |

Data presented as n/N (%) or median (IQR). \* p-value < 0.01 compared to controls. IQR = interquartile range, OC = oral contraceptive.

Table 2 shows the proportion of OC users in cases and controls stratified by tertile of metaGRS. The association of OC use with stroke was highest among women with the highest genetic risk of stroke (OR = 3.9; 95% C.I. 1.3-11.8) and lowest among women with the lowest genetic risk of stroke (OR = 1.5; 95% C.I. 0.6-3.9). Although not statistically significant, the formal test of interaction was consistent with our hypothesis (p = 0.07) that the genomic risk score modifies the association of OC use with ischemic stroke.

**Table 2.** Odds ratio for oral contraceptive use stratified by genetic risk score

| Tertile based on metaGRS | OR for OC use (95% C.I.) * | N OC users / N cases | N OC users / N controls |
| --- | --- | --- | --- |
| Upper Tertile | 3.9 (1.3 - 11.8) | 13 / 51 (25%) | 5 / 62 (8%) |
| Middle Tertile | 2.9 (1.0 - 8.0) | 12 / 47 (25%) | 7 / 66 (10%) |
| Lower Tertile | 1.5 (0.6 - 3.9) | 11 / 45 (24%) | 12 / 69 (17%) |
| All participants | 2.4 (1.4 - 4.3) | 36 / 143 (25%) | 24 / 197 (12%) |

\* p-value for metaGRS × OC use interaction = 0.07 (see text). OR = odds ratio, OC = oral contraceptive, C.I. = confidence interval.

## Discussion

In our cohort, OC use was significantly associated with the risk of ischemic stroke, as has been reported previously.^1–3^ Importantly, the odds ratio for ischemic stroke associated with OC-use was highest in the tertile with highest genomic risk of ischemic stroke. The formal test of interaction, although not statistically significant, is consistent with our hypothesis that the genomic risk score modifies the association of OC use with ischemic stroke.

OCs are the most used reversible method of contraception and comprise >20% of all contraception in current use in the U.S.. ^4^ Although ischemic strokes in OC users are rare, the high number of users highlights the importance of reducing excess risk.

Risk factors for OC use related ischemic stroke include older age, migraine (especially with aura), smoking, hypertension, and obesity.^4^ While many of the cardiovascular risk factors such as hypertension and diabetes often emerge by increasing age, the risk for ischemic stroke is suggested to be highest shortly after initiation of OCs.^2^ If a genomic risk score could identify women at latent risk, this could help to reduce the burden of early onset ischemic stroke and associated socioeconomic consequences.

The strength of our study is the novel study approach for estimating OC-associated ischemic stroke risk. The advantage of the metaGRS is that it takes into account genetic predisposition to several risk factors that are known also to increase OC-associated ischemic stroke risk such as smoking, hypertension and obesity (body mass index).^7^ We thus anticipate that it has better predictive power for OC associated ischemic stroke risk than individual GRSs.

Our analysis has several limitations. First, the most critical limitation lies in the small sample size. Larger sample size will enable analyses with both more metaGRS strata and tighter confidence intervals. Second, our study population is restricted to women of European ancestry from a small geographical region. Extending analyses into more diverse populations is important. This will be possible once a metaGRS based on other ancestries is available. Third, the current metaGRS does not include a polygenic risk score for deep venous thrombosis. We have previously shown that polygenic risk for deep venous thrombosis is associated with early-onset ischemic stroke at more significant level compared to later onset ischemic stroke, emphasizing that thromboembolic mechanisms likely play a more central role in early-onset ischemic stroke compared to later onset ischemic stroke.^6^ Considering OC’s impact on both venous and arterial thrombosis, adding a venous thrombosis polygenic risk into the model may further improve risk stratification. Lastly, future analyses should also determine the added value of the metaGRS over existing risk factor information.

Taken together, the results of our exploratory analysis highlight the need for international collaboration to generate sufficient sample sizes to determine whether a genomic risk score could be clinically useful in reducing OC-associated ischemic stroke risk. If a genomic risk score could provide improved identification of the subset of women with substantially increased risk of OC-associated ischemic stroke, this could influence prescribing guidelines and reduce stroke events.

## Supporting information

STROBE Checklist

## Non-standard Abbreviations and Acronyms

GEOS: The Genetics of Early-Onset Stroke Study
GRS: Genetic risk score
OC: Oral contraceptive

## References

1. Tepper NK, Whiteman MK, Zapata LB, Marchbanks PA, Curtis KM. Safety of hormonal contraceptives among women with migraine: A systematic review. Contraception. 2016;94:630–640.

2. Johansson T, Fowler P, Ek WE, Skalkidou A, Karlsson T, Johansson Å. Oral Contraceptives, Hormone Replacement Therapy, and Stroke Risk. Stroke. 0:10.1161/STROKEAHA.121.038659.

3. Lidegaard Ø, Løkkegaard E, Jensen A, Skovlund CW, Keiding N. Thrombotic Stroke and Myocardial Infarction with Hormonal Contraception. N. Engl. J. Med. 2012;366:2257–2266.

4. Teal S, Edelman A. Contraception Selection, Effectiveness, and Adverse Effects: A Review. JAMA. 2021;326:2507–2518.

5. Cheng Y-C, O’Connell JR, Cole JW, Stine OC, Dueker N, McArdle PF, Sparks MJ, Shen J, Laurie CC, Nelson S, et al. Genome-Wide Association Analysis of Ischemic Stroke in Young Adults. G3 GenesGenomesGenetics. 2011;1:505–514.

6. Jaworek T, Xu H, Gaynor BJ, Cole JW, Rannikmae K, Stanne TM, Tomppo L, Abedi V, Amouyel P, Armstrong ND, et al. Contribution of Common Genetic Variants to Risk of Early Onset Ischemic Stroke. Neurology [Internet]. 2022;Available from: https://n.neurology.org/content/early/2022/08/31/WNL.0000000000201006

7. Abraham G, Malik R, Yonova-Doing E, Salim A, Wang T, Danesh J, Butterworth AS, Howson JMM, Inouye M, Dichgans M. Genomic risk score offers predictive performance comparable to clinical risk factors for ischaemic stroke. Nat. Commun. 2019;10:5819.

