## Supplementary material for "Genomic Risk Scores and Oral Contraceptive-Associated Ischemic Stroke Risk": STROBE Checklist

STROBE Statement—checklist of items that should be included in reports of observational studies

|  | Item No. | Recommendation | Page  No. | Relevant text from manuscript |
| --- | --- | --- | --- | --- |
| **Title and abstract** | 1 | (*a*) Indicate the study’s design with a commonly used term in the title or the abstract | 2 | We performed a case-control |
|  |  | (*b*) Provide in the abstract an informative and balanced summary of what was done and what was found | 2 | Using a previously validated genetic risk score (metaGRS) for ischemic stroke based on 19 polygenic risk scores of vascular events and risk factors, we stratified our sample into tertiles of genomic risk. We evaluated the association between OC use and ischemic stroke within each tertile. We tested if the association between OC use and ischemic stroke depended on the genomic risk of stroke using logistic regression with an OC use × metaGRS interaction term. Among all women, OC use was significantly associated with ischemic stroke (odds ratio = 2.4, p = 0.002). The odds ratio for ischemic stroke associated with OC use decreased from 3.9 in the tertile with the highest genomic risk of ischemic stroke to 1.5 in the tertile with the lowest genomic risk. The formal test of interaction was consistent with our hypothesis (p = 0.07) that the genomic risk score modifies the association of OC use with ischemic stroke. |
| Introduction | | | |  |
| Background/rationale | 2 | Explain the scientific background and rationale for the investigation being reported | 4 | Despite several factors that increase OC-associated ischemic stroke have been identified, including age, smoking, and hypertension, 4 the exact mechanisms remain insufficiently understood. Identifying those women at risk is important to reduce the OC-associated risk of ischemic stroke. |
| Objectives | 3 | State specific objectives, including any prespecified hypotheses | 4 | The aim of our study was to evaluate whether genetic predisposition for ischemic stroke, measured by a validated genomic risk score, modifies OC-associated ischemic stroke risk. |
| Methods | | | |  |
| Study design | 4 | Present key elements of study design early in the paper | 4 | The Genetics of Early Onset Stroke (GEOS) Study is a population-based case-control study designed to identify genetic determinants of early-onset ischemic stroke and to characterize interactions with environmental risk factors. |
| Setting | 5 | Describe the setting, locations, and relevant dates, including periods of recruitment, exposure, follow-up, and data collection | 4 | Stroke cases aged 15-49 years old at the time of ischemic stroke were recruited from the greater Baltimore-Washington area over 4 time periods between 1992–2008, along with age and sex matched controls. Study subjects provided information from a standardized interview on age, ancestry, ethnicity, and history of hypertension, diabetes, and coronary artery disease. Current smoking status and OC use were defined use within 1 month before the event for cases and at a comparable reference time for controls. |
| Participants | 6 | (*a*) *Cohort study*—Give the eligibility criteria, and the sources and methods of selection of participants. Describe methods of follow-up  *Case-control study*—Give the eligibility criteria, and the sources and methods of case ascertainment and control selection. Give the rationale for the choice of cases and controls  *Cross-sectional study*—Give the eligibility criteria, and the sources and methods of selection of participants | 4 | Stroke cases aged 15-49 years old at the time of ischemic stroke were recruited from the greater Baltimore-Washington area over 4 time periods between 1992–2008, along with age and sex matched controls. |
|  |  | (*b*) *Cohort study*—For matched studies, give matching criteria and number of exposed and unexposed  *Case-control study*—For matched studies, give matching criteria and the number of controls per case |  |  |
| Variables | 7 | Clearly define all outcomes, exposures, predictors, potential confounders, and effect modifiers. Give diagnostic criteria, if applicable | 4-5 | Study subjects were genotyped with Illumina 1M array and imputed using the TOPMed reference panel on the University of Michigan Imputation Server, as previously described.6  We used a previously validated genomic risk score (metaGRS) for ischemic stroke that is based on 19 polygenic risk scores for vascular events and cardiovascular risk factors and validated in nearly 400,000 subjects from the UK BioBank. 7 We first stratified our sample of GEOS women into tertiles of genomic risk based on their metaGRS scores and evaluated the association between OC use and ischemic stroke within each tertile. We then tested if the association between OC use and ischemic stroke depended on genomic risk of stroke using logistic regression with an OC use × metaGRS interaction term. |
| Data sources/ measurement | 8* | For each variable of interest, give sources of data and details of methods of assessment (measurement). Describe comparability of assessment methods if there is more than one group | 4 | Page 5: Study subjects provided information from a standardized interview on age, history of hypertension, diabetes, and coronary artery disease. Current smoking status and OC use were defined use within 1 month before the event for cases and at a comparable reference time for controls.  Study subjects were genotyped with Illumina 1M array and imputed using the TOPMed reference panel on the University of Michigan Imputation Server, as previously described.6 |
| Bias | 9 | Describe any efforts to address potential sources of bias | NA |  |
| Study size | 10 | Explain how the study size was arrived at | NA |  |

| Quantitative variables | 11 | Explain how quantitative variables were handled in the analyses. If applicable, describe which groupings were chosen and why | NA |  |
| --- | --- | --- | --- | --- |
| Statistical methods | 12 | (a) Describe all statistical methods, including those used to control for confounding | 5 | We used a previously validated genomic risk score (metaGRS) for ischemic stroke that is based on 19 polygenic risk scores for vascular events and cardiovascular risk factors and validated in nearly 400,000 subjects from the UK BioBank. 7 We first stratified our sample of GEOS women into tertiles of genomic risk based on their metaGRS scores and evaluated the association between OC use and ischemic stroke within each tertile. |
|  |  | (*b*) Describe any methods used to examine subgroups and interactions | 5 | We then tested if the association between OC use and ischemic stroke depended on genomic risk of stroke using logistic regression with an OC use × metaGRS interaction term. |
|  |  | (*c*) Explain how missing data were addressed | NA |  |
|  |  | (*d*) *Cohort study*—If applicable, explain how loss to follow-up was addressed  *Case-control study*—If applicable, explain how matching of cases and controls was addressed  *Cross-sectional study*—If applicable, describe analytical methods taking account of sampling strategy | 4 | Stroke cases aged 15-49 years old at the time of ischemic stroke were recruited from the greater Baltimore-Washington area over 4 time periods between 1992–2008, along with age and sex matched controls. |
|  |  | (*e*) Describe any sensitivity analyses |  |  |
| Results | | | | |
| Participants | 13* | (a) Report numbers of individuals at each stage of study—eg numbers potentially eligible, examined for eligibility, confirmed eligible, included in the study, completing follow-up, and analysed | 7 | Characteristics of the 143 cases and 197 controls are shown in Table 1. |
|  |  | (b) Give reasons for non-participation at each stage | NA |  |
|  |  | (c) Consider use of a flow diagram | NA |  |
| Descriptive data | 14* | (a) Give characteristics of study participants (eg demographic, clinical, social) and information on exposures and potential confounders | 5-6, 10 | Table 1. |
|  |  | (b) Indicate number of participants with missing data for each variable of interest | 5 | All included participants had OC data and genetic data. |
|  |  | (c) *Cohort study*—Summarise follow-up time (eg, average and total amount) | NA |  |
| Outcome data | 15* | *Cohort study*—Report numbers of outcome events or summary measures over time | *NA* |  |
|  |  | *Case-control study—*Report numbers in each exposure category, or summary measures of exposure | *11* | *Tables 2.* |
|  |  | *Cross-sectional study—*Report numbers of outcome events or summary measures | *NA* |  |
| Main results | 16 | (*a*) Give unadjusted estimates and, if applicable, confounder-adjusted estimates and their precision (eg, 95% confidence interval). Make clear which confounders were adjusted for and why they were included | 11 | Table 2. |
|  |  | (*b*) Report category boundaries when continuous variables were categorized | NA |  |
|  |  | (*c*) If relevant, consider translating estimates of relative risk into absolute risk for a meaningful time period | NA |  |

| Other analyses | 17 | Report other analyses done—eg analyses of subgroups and interactions, and sensitivity analyses | 6, 11 | Table 2. Page 7: the formal test of interaction was consistent with our hypothesis (p = 0.07) that the genomic risk score modifies the association of OC use with ischemic stroke. |
| --- | --- | --- | --- | --- |
| Discussion | | | | |
| Key results | 18 | Summarise key results with reference to study objectives | 6 | In our cohort, OC use was significantly associated with the risk of ischemic stroke, as has been reported previously. 1–3 Importantly, the odds ratio for ischemic stroke associated with OC-use was highest in the tertile with highest genomic risk of ischemic stroke. The formal test of interaction, although not statistically significant, supports our hypothesis that the genomic risk score modifies the association of OC use with ischemic stroke even though test did not reach level of statistical significance. |
| Limitations | 19 | Discuss limitations of the study, taking into account sources of potential bias or imprecision. Discuss both direction and magnitude of any potential bias | 7-8 | In our exploratory analysis, the most critical limitation lies in the small sample size. However, our suggestive findings point to the need to increase sample size through international collaboration. Our cohort comes from a small geographical region and represents European ancestry and extending analyses into more diverse populations is important. The metaGRS has limitations in its application to early-onset ischemic stroke cohorts. We have previously shown that polygenic risk for deep venous thrombosis is associated with early-onset ischemic stroke at more significant level compared to later onset ischemic stroke, emphasizing that thromboembolic mechanisms likely play a more central role in early-onset ischemic stroke compared to later onset ischemic stroke.6 Considering OC’s impact on both venous and arterial thrombosis, adding a venous thrombosis polygenic risk into the model may further improve risk stratification. Larger sample size will also enable analyses with both more metaGRS strata and tighter confidence intervals. Future analyses should also determine the added value of the metaGRS over existing risk factor information and across different subtypes of ischemic stroke. |
| Interpretation | 20 | Give a cautious overall interpretation of results considering objectives, limitations, multiplicity of analyses, results from similar studies, and other relevant evidence | 8 | the results of our exploratory analysis highlight the need for international collaboration to generate sufficient sample sizes to determine whether a genomic risk score could be clinically useful in reducing OC-associated ischemic stroke risk. If a genomic risk score could identify a subset of women with substantially increased risk of OC-associated ischemic stroke, this could influence prescribing guidelines and reduce stroke events. |
| Generalisability | 21 | Discuss the generalisability (external validity) of the study results | NA |  |
| Other information | |  | | |
| Funding | 22 | Give the source of funding and the role of the funders for the present study and, if applicable, for the original study on which the present article is based | 8 | The study is supported by NIH R01NS086905, R01NS100178, R01NS105150. |

*Give information separately for cases and controls in case-control studies and, if applicable, for exposed and unexposed groups in cohort and cross-sectional studies.

**Note:** An Explanation and Elaboration article discusses each checklist item and gives methodological background and published examples of transparent reporting. The STROBE checklist is best used in conjunction with this article (freely available on the Web sites of PLoS Medicine at http://www.plosmedicine.org/, Annals of Internal Medicine at http://www.annals.org/, and Epidemiology at http://www.epidem.com/). Information on the STROBE Initiative is available at www.strobe-statement.org.
